# Interaction of citrate-stabilized gold nanoparticles with umbilical cord RBC

**DOI:** 10.1101/2025.06.07.25329131

**Authors:** Sarika Hinge, Gauri Kulkarni

## Abstract

**Introduction:** Umbilical cord blood contains stem cells which are useful in chemotherapy, implantation; however, cord RBC is considered as a waste product. It is necessary to use the umbilical cord blood for neonatal blood transfusion to avoid diseases. Nowadays, nanoparticles are used in biomedical and pharmaceutical industry. Nanoparticles interact with blood via different routes. In addition, these nanoparticles used in biomedicine, drug delivery, bioimaging, and to increase the shelf life of RBC. Nevertheless, research of nanoparticle interaction with umbilical cord red blood cells is still insufficient. Thus, in the present study, interaction between umbilical cord blood-RBC and citrate-stabilized gold nanoparticles is studied.

**Methods:** Umbilical cord rbc isolated from whole blood immediately after the collection using centrifuge. PBS buffer is used as a physiological media and washing reagent. After, interaction of isolated RBC (2% suspension) with different concentration of citrate stabilized gold nanoparticle were investigated using UV-Visible spectroscopy, Raman spectroscopy, and Interfacial tension measurement technique.

**Results:** UV-Visible spectroscopy is used to investigate the changes in Soret band and oxygenation bands (Q1 and Q2). Variation in absorption intensity is observed in umbilical cord RBC incubated citrate stabilized gold nanoparticles. However, there is no significant shift in Soret band and Q1, Q2 band of cord-RBC with Au nanoparticles. In case of Raman spectroscopy, enhancement in disulphide band region (526 cm^-1^ and 579 cm^-1^) and change in the spin state marker is investigated. While, interfacial tension decreases for 10 µl and increases for 100 µl and 200µl of 40 nM AuNP.

**Conclusion:** The umbilical cord rbc remains unaffected by 10 µl of citrate-stabilized gold nanoparticles (40 nM).

## Introduction

Umbilical cord blood RBC differs from adult RBC in many aspects and has a greater affinity for oxygen. Cord RBC shows distinguishing characteristics such as cell membrane composition, morphological structure, surface tension, viscosity, pH, haemoglobin concentration, and oxygen binding affinity [1],[2]. Generally, cord RBCs are considered as a waste product but to some extent they can be used for blood transfusion in neonates [3]. Adult RBC transfusion in the neonate may lead to various diseases related to respiratory, lung, and eye. Therefore, it is necessary to store cord RBCs for a long duration due to their unique features and small volume availability. Umbilical cord blood contains stem cells which can produce other types of cells. These cells are used for transplantation in cancer therapy [4]. Cord RBCs have a short life span of 45-70 days as compared to adult RBCs of 120 days. The large size of cord RBC with more cell rigidity makes the cell less deformable within the small blood vessels. Cord blood-RBC contains more lipids, phosphorous, and cholesterol per cell than adults [5]. Cord RBC shows a decrease in sedimentation rate and roulex formation. Osmotic fragility of cord RBC is lower as compared with adult, due to pH values. The concentration of Haemoglobin in the cord (15.7-17.9 g/dm) is significantly higher than that of Adults [6],[7]. Cord blood contains different Haemoglobin i.e. HbA, HbF, and HbA2. Fetal haemoglobin (HbF) has a higher affinity to bind to oxygen than adults. Cord RBC membrane becomes less dielectric as compared with adult RBC when measured by the dielectrophoresis technique. Cord blood has high-level proteins, K, Ca, P, and folic acid. Cord blood contains hematopoietic stem cells (HSC) which make every type of cell in the blood. Transplants of HSC from cord blood are used to treat blood diseases like leukemia. RBCs from cord blood contain fetal Hb which is dominant in newborns, therefore cord RBCs are more suitable for neonatal transfusion. Cord RBC is less deformable than adult RBC due to lower elastic compressibility, structural, and permeability differences. Cord RBCs are more prone to mechanical stress than adult cells [11]. Cord RBCs have a high percentage of cholesterol and phospholipids with lower membrane fluidity. Lipid composition is important in part of the physical properties of cord-RBC membranes [12].

Cord-RBC is major parameter to understand the health status of newborn baby and mother [15]. During pregnancy, health of the mother plays important role in blood rheology of baby. Pathophysiological investigation of Cord RBC like RBC aggregation, sedimentation rate provides information whether the mother suffered from hypoxia, hypertension, thalassemia, etc. Low viscosity of plasma and less proteins leads to RBC aggregation in cord blood [16], [17]. Cord RBC contains more membrane-bound Haemoglobin than adults. Fetal Haemoglobin is more susceptible to oxidation stress than HbA and protects proteins against the stress [18]. Iron oxide nanoparticles (SPION) and cord blood-derived natural killer cells are used for cancer cell therapy [3]. Targeted transplantation of cord blood with immunomagnetic nanoparticles was used to repair corneal defects [8]. Stem cells present in the umbilical cord RBC are used in the defense mechanism of lymphoma, and leukemia. Recently, it has been reported that damaged stem cells due to certain medical reasons can be repaired using nanoparticles and drugs [9]. Some nanoparticles like silica-coated magnetic nanoparticles are used to label images. Ektacytometry, UV-visible spectroscopy, Blood Gas analysis, FTIR, Mossbauer spectroscopy, Raman spectroscopy, Dynamic Light Scattering, AFM, SNOM Micropipette aspiration techniques are used to understand the biophysical state of RBC. Very few reports are available on umbilical cord RBC-nanoparticle interactions. Citrate stabilized gold nanoparticles were used in labelling, immobilization of protein, biosensing, and targeted drug delivery application. In the present work, umbilical cord RBC-citrate stabilized gold nanoparticle interactions are investigated using salinity test, UV-visible spectroscopy, Raman spectroscopy, and Interfacial tension measurement techniques.

## Materials and Methods

### Chemicals

Gold nanoparticles (5 nm) were procured from Sigma Aldrich; the solution is stabilized by using Citrate buffer with O.D. 1., phosphate buffer saline.

### Cord blood Collection

Cord blood was immediately collected after delivery in a standard hospital under the supervision of a medical expert and stored in an EDTA vial. Ethical approval for this study was obtained from Savitribai Phule Pune University ethical committee with the consent of subject **(Ref. No. SPPU/IEC/2021/129)**.

Cord blood was collected from standard hospital under the supervision of gynecologist and store in EDTA vial. Cord red blood cells were isolated from whole blood using centrifugation for 10 minutes at 25 °C at 3000 rpm. RBCs were washed with PBS buffer saline pH-7.34 for three times. For experiment purposes, 2% suspensions of Cord-RBC were prepared with PBS as a diluting reagent. All experiments performed with Cord-RBC were conducted in accordance with the rule of Institutional ethical committee. Ethical approval **(Ref. No. SPPU/IEC/2021/129**) for this study was obtained from Savitribai Phule Pune University ethical committee with subject consent.

### Umbilical cord blood RBC-citrate stabilized gold nanoparticles

Cord RBCs were treated with citrate-stabilized gold nanoparticles with concentrations of 10 µl and 100 µl of 40 nM for 12 hr of incubation at 37 LC. Characterization was performed using salinity test, UV-visible spectroscopy, Raman Spectroscopy, and Interfacial tension measurement techniques.

## Results and Discussions

### UV-visible spectroscopic study of citrate stabilized Au nanoparticle interaction with Cord blood-RBC

Absorption spectra of Cord RBC-Au nanoparticle interactions were recorded using a UV-visible spectrophotometer (JASCO - 760). UV-visible spectra measurements were performed using 2% suspension of cord RBC and RBC incubated with citrate stabilized gold nanoparticles for different volumes i.e. 10 µl, and 100 µl of 40 nM in PBS buffer. Absorption of the haem group inside the RBC and its associated oxygenation bands i.e. Q1 and Q2 were examined in the present study.

### Raman spectroscopic study of Au nanoparticle interaction with Cord-RBC

Raman spectra of cord RBC and citrate-stabilized gold nanoparticles were recorded using a Renishaw spectrophotometer (inVia) which is shown in **Figure 2**. RBC was incubated with 40 nM concentration of Au nanoparticles for 4 hr. The laser power delivered to the sample was 15 mW to avoid photodamage. One drop of 15µl Cord - RBC and citrate stabilized Gold nanoparticle-incubated cord-RBC was drop cast on the Aluminum substrate and samples were settled for 15 min.

**Figure 1:**
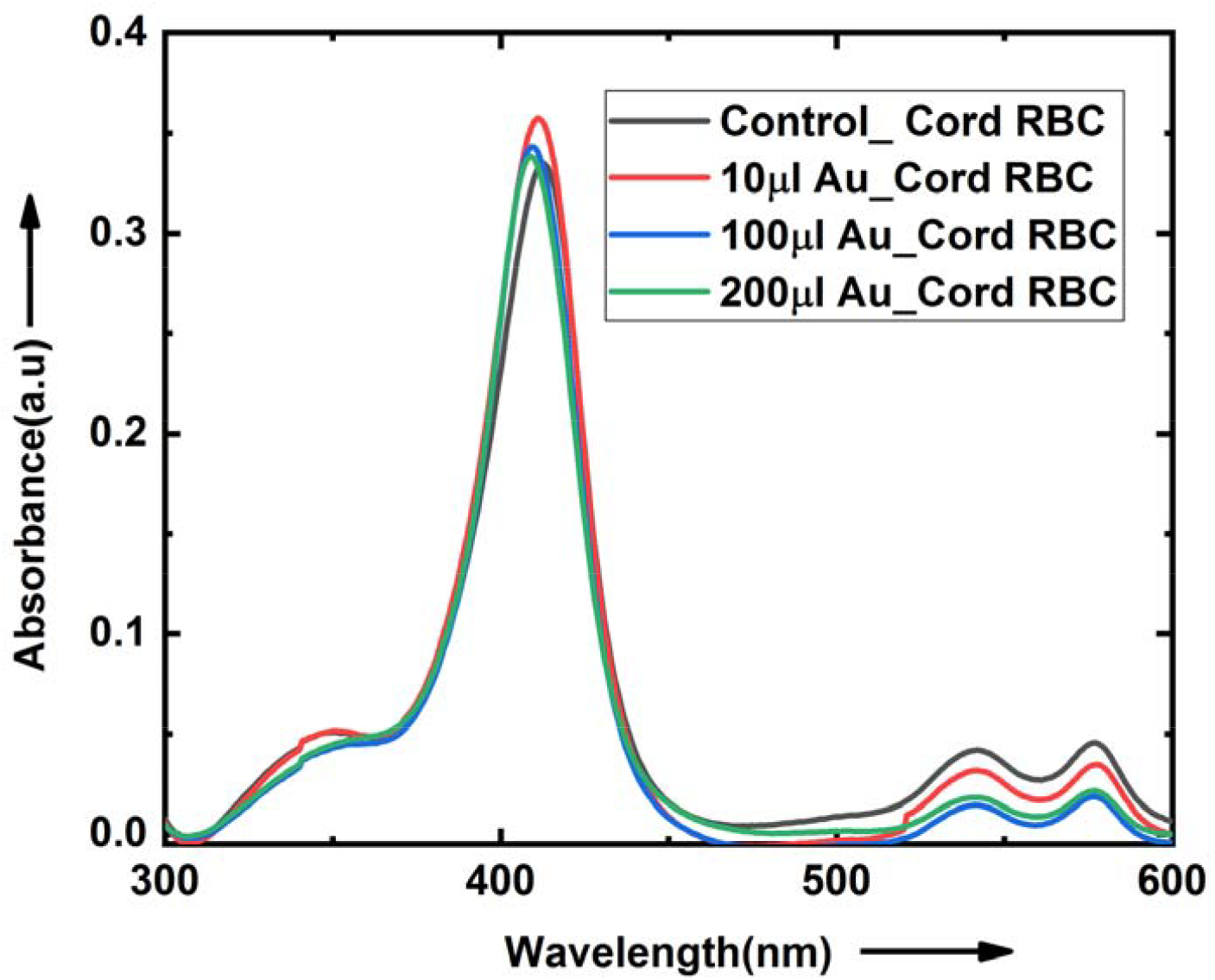
UV-visible spectroscopy of Au nanoparticles treated Cord RBC. UV-visible absorption spectroscopy of Adult RBC and Cord RBC shows peaks at 281.53nm, 347.33nm, 420.51nm (Soret band), 543.77nm (Q1) and 578.11nm (Q2) which are shown in **Figure 1**. UV-Visible spectra of umbilical cord rbc shows Soret band peak position at 420 nm. Similarly, the Q1 and Q2 peaks representing oxygenation state at 543 nm and 578 nm, respectively. Variation in absorption intensity is observed in umbilical cord rbc incubated citrate stabilized gold nanoparticles **(see Table S1)**. However, there is no significant shift in Soret band and Q1, Q2 band of cord-RBC with Au nanoparticles. Salinity test of cord RBC for different molar concentration of NaCl is provided in Suppl. **Figure S1**.

**Figure 2.**
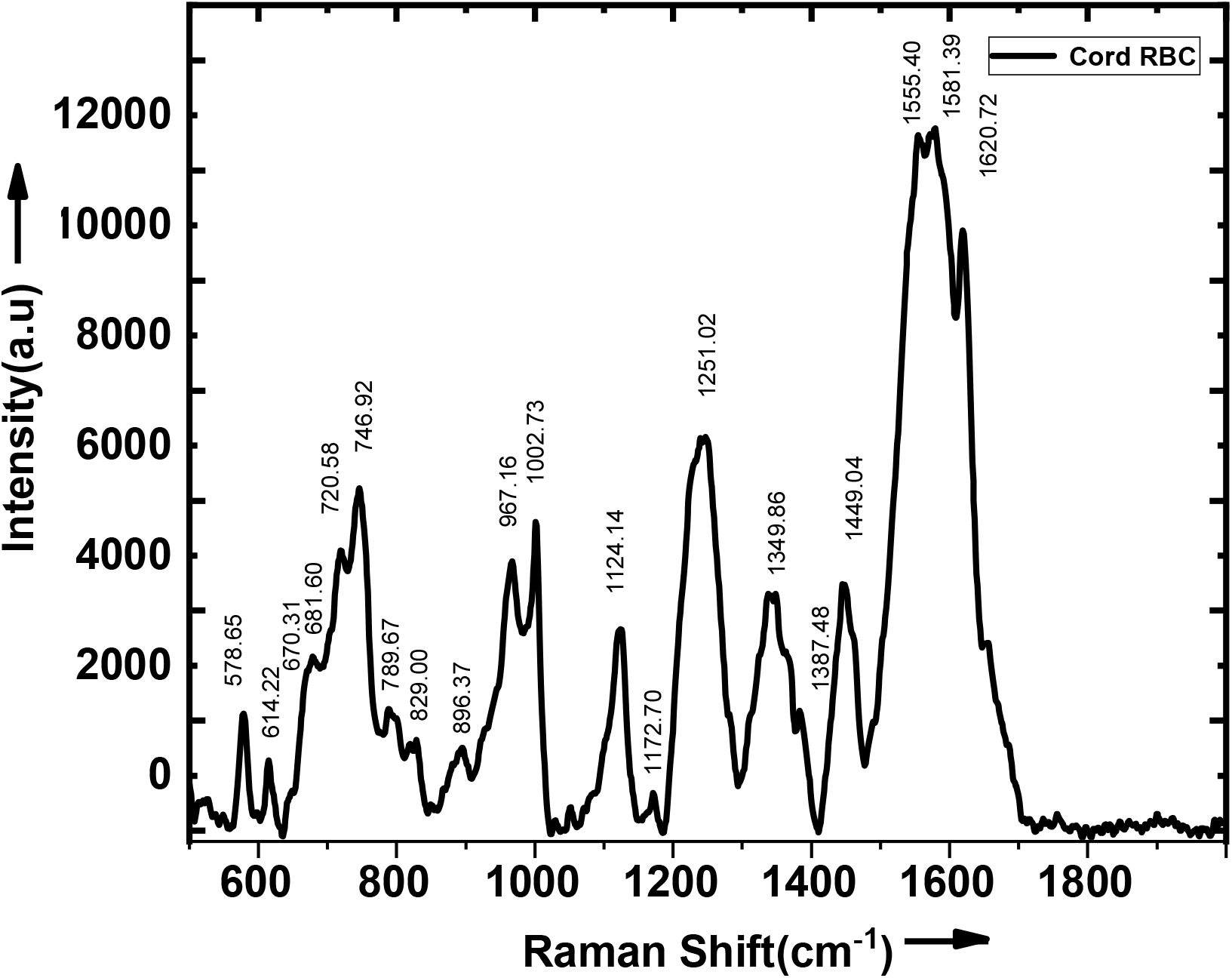
Raman spectrum of isolated Cord RBC from whole Cord blood

Raman peak and its signature associated with umbilical cord -RBC is as follows:

1. Spin state marker region is in between 1500-1640 cm^-1^ appearing at 1550 cm^-1^, 1581 cm^-1^, and 1620 cm^-1^.
2. Pyrrole ring vibration modes of Haemoglobin are observed at 1387 cm^-1^, 1371 cm^-1^, 1349 cm^-1^ which corresponds to half ring symmetric stretch.
3. Ring deformation is observed at 670 cm^-1^ and 849 cm^-1^ is assigned to single-mode stretching vibrations for the amino acids and valine polysaccharides.
4. The peak position at 720 cm^-1^, 968 cm^-1^, 1738 cm^-1^ corresponds to lipid assignment,1125 cm^-1^ is appearing for ν(C=C) Acyl backbone in lipid.
5. The peak associated with 614 cm^-1^ is for cholesterol ester, Nucleic acid is appearing at 829 cm^-1^, 1340 cm^-1^
6. Sulphide stretching is observed in protein at 526 cm^-1^, phosphatidylserine and phosphodiester is appearing at 579 cm^-1^ and 896 cm^-1^, Cytosine, guanine and tyrosine assignment is at 1173 cm^-1^ and 1248 cm^-1^.
7. C-O stretching, C-N stretching of protein is observed at 1053 cm^-1^, CH_3_ band at 1384 cm^-1^, C=O at 1717 cm^-1^ and 1759 cm^-1^ is assigned for C=O stretching.

The detail information related to band assignment is provided in **Table S2**.

### Raman spectra of Cord RBC incubated with Gold nanoparticles

Enhancement of peak intensity at frequency position 526 cm^-1^ and 579 cm^-1^ indicates that bridges of disulfide region increase in gold nanoparticles treated cord RBC. This result is consistent with bond formation between protein-glutathione and sulfide bridge, indicating oxidative stress. At the spin marker region, the CH_3_ band disappears at citrate-stabilized gold nanoparticles incubated RBC as shown in **Figure 3**.

**Figure 3.**
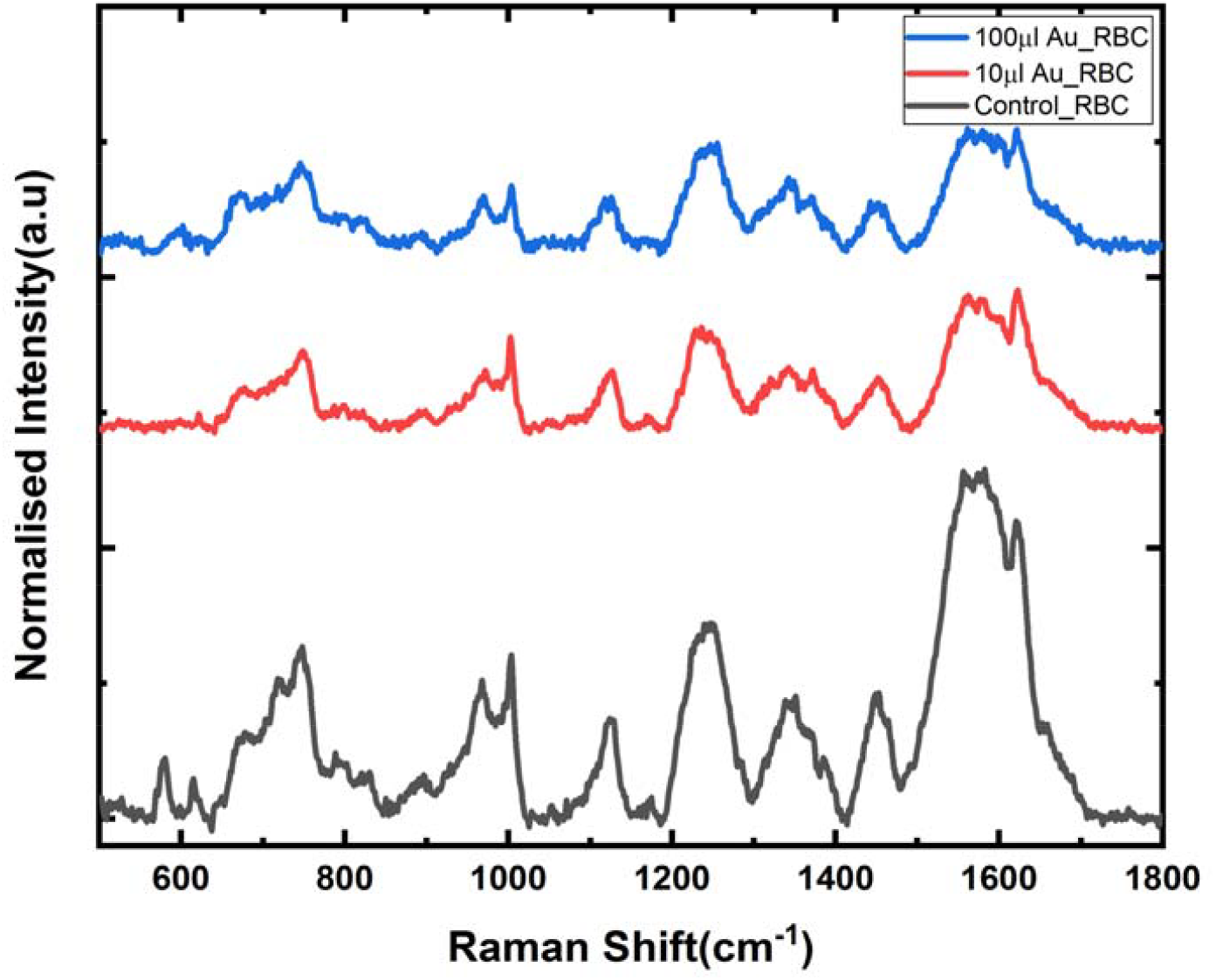
Raman spectra of Cord RBC incubated with Gold nanoparticles with different concentrations a) control b) 5 µl c)10 µl d)100 µl

### Interfacial tension measurement of Cord RBC suspension using the Pendant drop method

Interfacial tension of umbilical cord RBC was measured using SCA 20 software and the Pendant drop method. Images of umbilical cord RBCs were extracted and fitted with Young’s Laplace Equation.

Interfacial tension of Cord-RBC treated with citrate stabilized gold nanoparticle of volume 10 µl from 40 nM concentration has a lower value as compared with pristine cord-RBC as shown in **Figure 4**. However, Cord RBC treated with higher concentration i.e. 100 µl and 200 µl of 40 nM gold nanoparticles has high interfacial tension. The IFT of umbilical cord RBC influences the pathophysiology of surrounding medium which depend on the physical state of the RBC membrane.

**Figure 4.**
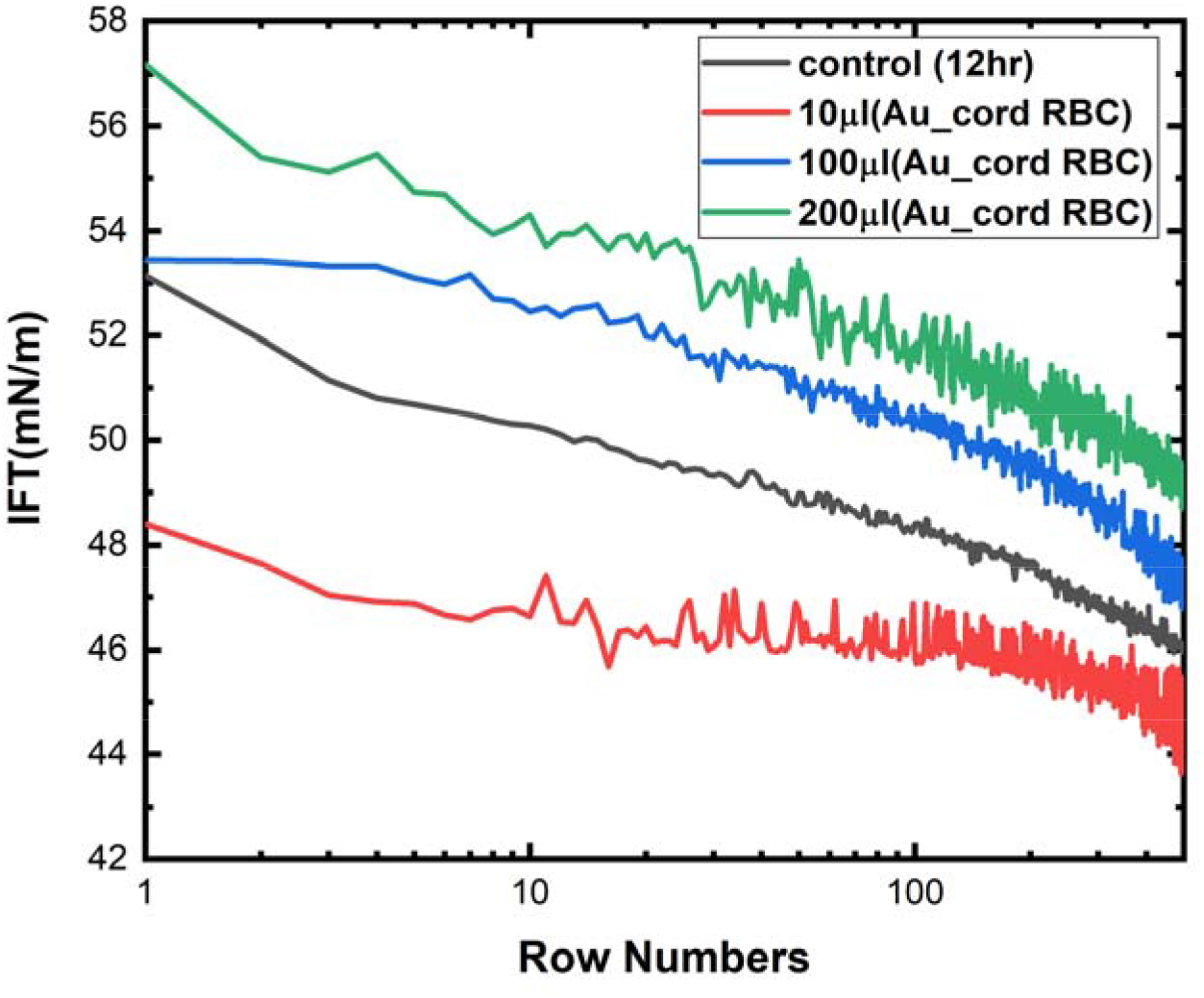
Interfacial tension Measurement of Cord Blood-RBC incubated with gold nanoparticles a) control b)5 µl c)10 µl d)100 µl e)200 µl

## Conclusion

Variations in umbilical cord RBC were observed in the presence of low and high concentration of citrate stabilized gold nanoparticles viz.10 µl and 100 µl of 40 nM. Reduction in signal intensity is observed in UV-visible and Raman spectroscopy.

Furthermore, with a high concentration (100 µl), the interfacial tension was increased and reduced for 10 µl of citrate-stabilized gold nanoparticles.

## Author Contributions

Sarika Hinge: conception, design, data collection, Data analysis, results interpretation, validation, and manuscript preparation. Gauri Kulkarni: Validation and supervision. All authors reviewed the Results and approved the final version of the Manuscript.

## Data Availability

All data generated or analyzed during this study are included in this published article.

## Ethics Declarations

### Consent to participate

Not applicable.

## Ethical approval

Not applicable.

